# A pilot study to investigate the fecal dissemination of SARS-CoV-2 virus genome in COVID-19 patients in Odisha, India

**DOI:** 10.1101/2020.05.26.20113167

**Authors:** Shantibhusan Senapati, Jaya Singh Kshatri, Punit Prasad, Jyotirmayee Turuk, Sanghamitra Pati, Ajay Parida

**Author notes:** These authors contributed equally (Shantibhusan Senapati, Jaya Singh Kshatri, Punit Prasad). **Address for correspondence:** Dr. Ajay Parida, FNASc, FNAAS, Or, Dr. Sanghamitra Pati, MBBS, MD, MPH.

## Abstract

In infectious diseases, the routes of transmission play major roles in determining the rate and extent of disease spread. Though fomites and aerosol droplets are major sources of SARS-CoV-2 human to human transmission, studies have also reported possible involvement of other routes of transmission like fecal-oral. Multiple studies around the world have reported shedding of the SARS-CoV-2 viral genome in certain COVID-19 patient fecal samples. Hence, the major objective of this study was to get the experimental evidence whether in Indian COVID-19 patients fecal dissemination of the SARS-CoV-2 genome occurs or not. Information obtained from twelve number of patients from a COVID-19 hospital of Odisha has demonstrated that both symptomatic and asymptomatic Indian patients could be positive for the SARS-CoV-2 genome in their fecal component. The findings have also established a protocol to collect and extract viral RNA for SARS-CoV-2 detection in fecal samples. Together, the study supports the hypothesis of possible fecal-oral transmission of SARS-CoV-2 virus and provides a rationale to extend this study in a larger cohort of patient samples and correlate the significance of the SARS-CoV-2 virus genome detection in fecal samples with disease severity and transmission.

## Introduction

Recently, the novel severe acute respiratory syndrome coronavirus, SARS-CoV-2 has spread across the globe, including all states of India. The most common human-to-human transmission mode for this virus is through fomites, physical contact, aerosol droplets, nosocomial transmission. Recently, a number of studies from different parts of the world have reported shedding of the SARS-CoV-2 genome in the fecal sample of coronavirus disease 2019 (COVID 19) patients (Chen et al., 2020a; Chen et al., 2020b; Wu et al., 2020). Interestingly, a few studies have reported the presence of SARS-CoV2 in stool samples when nasopharyngeal samples were negative in a substantial number of patients (Chen et al., 2020a; Wu et al., 2020). Findings of gastrointestinal symptoms like diarrhea in certain COVID-19 patients suggest a possible direct effect of the SARS-CoV-2 virus in enteric infection (Huang et al., 2020). Experimental confirmation of Angiotensin-converting enzyme 2 (ACE2) as the entry receptor for SARS-CoV-2 virus (Walls et al., 2020; Wang et al., 2020), and high level of its expression in enterocytes provided a rationale to hypothesize that in addition to lung epithelial cells, enterocytes might be another type of target cells for this virus (Chen et al., 2020b; Liang et al., 2020). Moreover, it has been reported that human enterocytes do get infected by the SARS-CoV-2 virus and these cells support its replication (Lamers et al., 2020; Xiao et al., 2020; Zang et al., 2020). Isolation of live SARS-CoV-2 virus from stool samples have also raised possibilities of fecal-oral transmission of this virus (Gu et al., 2020).

To date, the rate of COVID-19 spreading in India has not reached the predicted growth rate. Despite a large population size and improper hygienic practices, India have reported lesser incidence of infection compared with the western countries. A high proportion of asymptomatic cases and less mortality rate in the Indian population is also a striking difference compared with the other countries. These observations have provided an impetus for scientific communities to propose multiple hypotheses that might be playing roles in the aforementioned differential manifestation of COVID-19. It has been proposed that the genetic and immune makeup of the different human populations, environmental conditions, human-associated microbiota, food habits, etc. might be contributing to the spread and severity of this disease (Butler and Barrientos, 2020; Cao et al., 2020; Dhar and Mohanty, 2020; Wigginton and Boehm, 2020). It is also possible that all these factors might regulate the incidence of fecal shedding/ dissemination of infectious SARS-CoV-2 virus or its genome in the Indian population. Based on these observations, a pilot study was conucted to get an insight, whether fecal dissemination of the SARS-CoV2 genome also happens in the Indian population?

## Materials and methods

### Study design and patients

The study was approved by the Institutional human ethical committee, ILS, Bhubaneswar, and the Govt. of Odisha, India. Informed consent was obtained from all the patients who participated in this study. All the samples used in this study were collected from patients admitted to a COVID-19 hospital, Odisha. Before hospitalization, all patients were tested positive for SARS-CoV-2 RNA in nasopharyngeal swab specimens through real-time reverse polymerase chain reaction (RT-qPCR). The initial diagnosis was done by COVID-19 testing centers approved by the Indian Council of Medical Research (ICMR) at Odisha. Details of patient information including clinical data were obtained from the hospitals are listed in Table 1.

### Sample collection and RNA isolation

The rectal swab (RS) and corresponding nasopharyngeal (NPS) swab for all the patients were collected on the same day by an experienced clinician of the COVID-19 hospital. The sterile swabs were used to collect samples from rectum and nasopharyngeal in 3 ml of DNA/RNA shield buffer (Zymo Research) and 3 ml of the virus transport medium (VTM) (HiMedia) respectively. As per the product information sheet, the DNA/RNA shield buffer inactivates infectious agents including virus and also stabilize DNA/RNA in the samples. All the samples were transported and stored at 4°C. For viral RNA isolation, the samples were processed at BSL3 facility of ILS, Bhubaneswar, a recognized COVID-19 testing center. Viral RNA from RS and NPS samples was isolated using QIAamp Viral RNA Mini Kit (Qiagen). Before RNA isolation, MS2 phage control 10 μl from TaqPath™ COVID-19 multiplex real-time PCR diagnostic kit (Thermo Fisher Scientific) was added to the lysis buffer from QIAamp kit. MS2 phage control acts as an internal control for RNA isolation protocol. For isolation of RNA from RS samples, we optimized the protocol and all the samples were processed identically. Initially, all the samples were centrifuged for 5 min at 4000*g, and then 1.5 ml of the RS suspension were passed through a 0.22 micron filter. For each sample, 280 μl of the filtrate were processed for RNA isolation as per the QIAamp protocol. To isolate viral RNA from NPS, 140 μl of VTM from 3 ml was used for each sample. RNA elution for each sample was done with 50 μl of EB (Qiagen).

### Reverse polymerase chain reaction (RT-PCR)

The RT-qPCR assay was conducted following the current protocol followed by ILS for COVID-19 screening. TaqPath™ COVID-19 multiplex real-time PCR diagnostic kit (Thermo Fisher Scientific) was used to detect the presence of the viral genome. The multiplex TaqMan reaction detects three SARS-CoV2 genes, mainly N gene (VIC), S gene (ABY), ORF1ab (FAM) along with MS2 (JUN). All the RT-qPCR reactions are set along with the positive control that contains SARS-CoV2 genomic regions and non-template control (NTC). The positive control reaction verifies the RT-PCR reaction setup and reagent integrity while NTC control for cross-contamination during RNA isolation and reaction setup. The samples were run in Applied Biosystems QuantStudio 6 Flex Real-Time PCR System as per the manufacturer’s protocol and the data was analyzed using QuantStudio™ Real-Time PCR software (Thermo Fisher Scientific). The multicomponent plot for fluorescence intensity, amplification curve, and Ct values derived from the relative threshold was used to confirm positive and negative for SARS-CoV2 infection in patient samples.

## Results

In RT-qPCR based disease diagnosis, the stability of DNA or RNA is a crucial point to avoid false-negative results. In our protocol to isolate RNA from rectal swab samples, we use DNA/RNA shield buffer, which confirms the suitability of this reagent for the collection of biological samples for SARS-CoV-2 genome detection and stability of RNA. The use of this reagent might also inactivate infectious agents and thus reduce the risk of exposure to infectious viral particles during handling. The RT-qPCR data analysis from 12 RS and corresponding NPS samples showed amplification of SARS-CoV2 RNA in six stool samples and eleven NPS samples. The amplification curve and the Ct values for different genes are presented in Figure 1. Six samples from RS have Ct values that were not dertermined (ND), which may be due to low sample input or actual absence of viral genome in these samples. The corresponding NPS samples showed very high amplification curves (Fig 1 left panel). The positive and negative control reactions showed amplification was as expected (data not shown). Based on the available clinical data, since the time of first SARS-CoV2 detection to the time of sample collection, seven patients were symptomatic and five were asymptomatic (Table 1). Four out of 5 (80%) asymptomatic and 2 out of 7 (28%) symptomatic cases were tested positive for the presence of viral genome in the rectal swab samples, respectively. The summary of clinical symptoms and the medical history of all the patients are shown in Table 1. One patient (COVID19/GNG03) was found negative for both NPS and rectal swab samples, which indicates that the patient might have recovered from the disease during the period of hospitalizaion.

**Figure 1:**
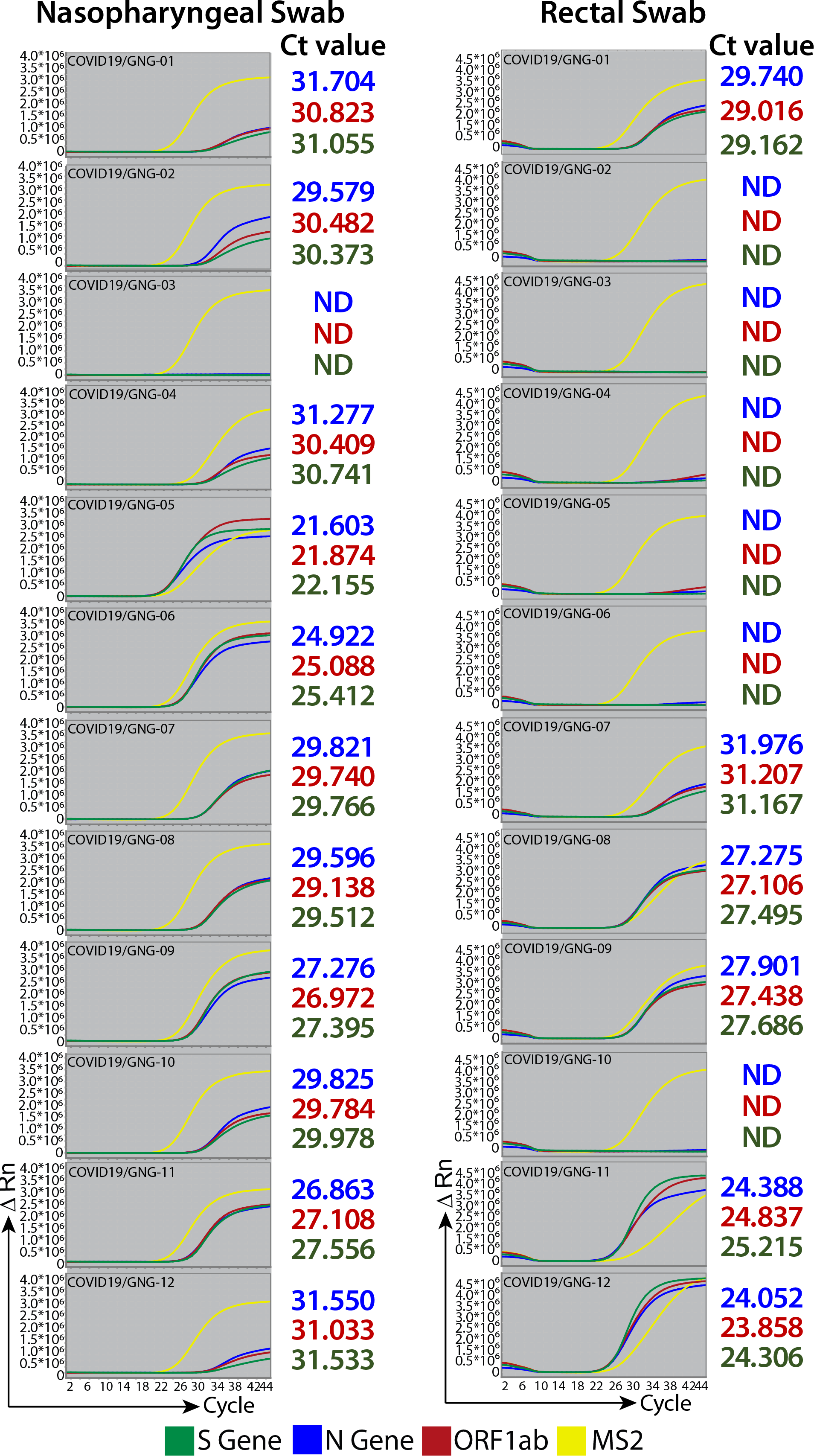
RT-qPCR profile of nasopharyngeal and rectal swabs from COVID-19 patients. The RT-qPCR amplification plots of nine COVID-19 patient’s nasopharyngeal (NPS) and rectal swabs (RS) are represented in the left and right panel respectively. The patient ID is mentioned as COVID19/GNG-01-12. The three viral genes amplification curves for N gene, ORF1ab, and S gene are labeled as blue, maroon, and green respectively while MS2 internal control is labeled with yellow. The amplification curves are plotted for Rn (Y-axis) vs cycle (X-axis). The Ct values for the respective RT-qPCR samples are shown alongside the graph.

**Table 1:** Comprehensive data for 12 COVID-19 patients from Odisha.

| **PATIENT ID**<br>(For this Study) | **AGE**<br>**RANGE** | **GENDER** | **COVID HOSPITAL**<br>**ADMISSION DATE** | **SAMPLE**<br>**COLLECTION DATE**<br>**FOR THE STUDY** | **CLINICAL STATUS**<br>**(FIRST**<br>**LABORATORY TEST)** | **CLINICAL STATUS**<br>**(DURING SAMPLE**<br>**COLLECTION)** |
| --- | --- | --- | --- | --- | --- | --- |
| COVID19/GNG-01 | 15-25 | Male | 10th May | 12th May | Symptomatic | Asymptomatic |
| COVID19/GNG-02 | 26-35 | Male | 10th May | 12th May | Symptomatic | Asymptomatic |
| COVID19/GNG-03 | 15-25 | Male | 04th May | 12th May | Symptomatic | Asymptomatic |
| COVID19/GNG-04 | 15-25 | Male | 10th May | 12th May | Symptomatic | Asymptomatic |
| COVID19/GNG-05 | 15-25 | Male | 11th May | 12th May | Asymptomatic | Symptomatic |
| COVID19/GNG-06 | 15-25 | Male | 11th May | 12th May | Asymptomatic | Symptomatic |
| COVID19/GNG-07 | 15-25 | Male | 06th May | 12th May | Asymptomatic | Asymptomatic |
| COVID19/GNG-08 | 15-25 | Male | 05th May | 12th May | Asymptomatic | Asymptomatic |
| COVID19/GNG-09 | 26-35 | Male | 10th May | 12th May | Asymptomatic | Asymptomatic |
| COVID19/GNG-10 | 36-45 | Male | 11th May | 12th May | Asymptomatic | Asymptomatic |
| COVID19/GNG-11 | 26-35 | Male | 11th May | 12th May | Asymptomatic | Asymptomatic |
| COVID19/GNG-12 | 15-25 | Male | 04th May | 12th May | Symptomatic | Symptomatic |

### Discussion and conclusion

To best of our knowledge this pilot study is the first report that confirms that in the eastern Indian COVID-19 patients, fecal dissemination of the SARS-CoV-2 genome is evident. Although the RT-qPCR results do not confirm the dissemination of infectious viral particles in the RS samples, our findings support the hypothesis of fecal-oral transmission of the SARS-CoV-2 virus (Heller et al., 2020). One study has already reported the isolation of live SARS-CoV2 viral particles from the fecal matter of patients suffering from COVID-19 (Gu et al., 2020). At the same time several studies have demonstrated that SARS-CoV-2 infects gastrointestinal epithelial cells and these cells support replication of this virus (Lamers et al., 2020; Xiao et al., 2020; Zang et al., 2020). Continuous detection of viral RNA even after the absence of the viral genome at the nasopharyngeal region indicates that infected enterocytes might be the source of these virions in the gastrointestinal tract. Although studies suggest that the harsh acidic environment of the stomach and alkaline condition of intestine neutralize these viruses (Zang et al., 2020), the chances of live virus shedding can’t be completely ruled out. Detection of viral nucleocapsid protein in rectal epithelial cells of COVID-19 patients suggests that some infectious viral particles might have escaped the gastrointestinal harsh environment (Xiao et al., 2020). Particularly in the case of patients with diarrhea, the bowel clearance might happen in such a short period that virus particles may not get effectively neutralized in the gut (Zang et al., 2020). Proton pump inhibitors (PPIs) are known to promote the survival of oral bacteria in the lower GI tract, and this is most likely through the acid-antisecretory effect of these drugs (Bruno et al., 2019). Similarly, studies have also shown that continuous PPI therapy is associated with an increased risk of enteric viral infection (Vilcu et al., 2019). Therefore, it is possible that in COVID-19 patients who might have been treated with any PPIs, the gastrointestinal neutralization of the virus might not be very effective.

Undoubtedly, earlier reports (Heller et al., 2020) and our findings warrant further indepth studies to understand the possible gastrointestinal route of this virus transmission. This study with a larger number of cohorts and multiple times of sampling will provide further information about the correlation between the dissemination of the viral genome with the severity of the disease in the Indian population. In the current scenario, optimum sanitary and hygienic measures should be adapted to avoid possible fecal-oral transmission of this virus. Importantly, hospitals might adapt a COVID-19 fecal sample screening before any planed gastrointestinal surgeries. The potential fecal-oral transmission should be taken into consideration to control the virus spread.

## Data Availability

All the data are given along with the manuscript. For any other information the corresponding author might be contacted.

## Acknowledgments

The study is a part of the research activities conducted by the Odisha COVOD-19 study group. The contribution of all the members of this group (supplementary information) is sincerely appreciated. The authors are also truly thankful to all the members of the COVID-19 screening group of ILS for their invaluable support. Especially we would like to thank Aliva P Minz, Swati Madhulika, Kautilya Kumar Jena, Satya Ranjan Sahu, Omprakash Shirivas, Sanchari Chatterjee, and Priyanka Mohapatra for their assistance. We also thank Dr. Ashish Sadangi, MKCG Medical College for his support for sample collection. We thank all the patients who participated in this study. The support provided by the Health Department of Odisha is highly appreciated. Financial support from the Department of Biotechnology & Indian Council of Medical Research, Govt. of India is greatly acknowledged.

